# Single center Automated, Multi-Source deeply Phenotyped Heart Transplant Registry as a template to build tailored data infrastructure

**DOI:** 10.64898/2026.02.07.26345785

**Authors:** Khush Patel, Todd N. Eager, Mark Ghobrial, Linda W Moore, Ashrith Guha, Cindy Martin, Mehmet H. Akay, Laurie Loza, Stephen L Jones, Ahmed O. Gaber, Arvind Bhimaraj

## Abstract

**Background:** Traditional heart transplant registries often lack the granularity required for deep phenotyping and rely on labor-intensive manual abstraction. We describe the methodology and validation of a next-generation, automated, multi-source registry designed to address these limitations.

**Methods:** Utilizing a High-Performance Computing environment, we integrated structured data from Epic data warehouses (Clarity and Caboodle), external molecular diagnostics, and verified UNOS survival records. A custom deterministic rule-based Natural Language Processing (NLP) engine was developed to extract echocardiographic measures, rejection grades, and vasculopathy scores from over 21,000 unstructured clinical reports.

**Results:** The Houston Methodist J.C. Walter Jr. Transplant Center Precision Registry and Platform-Heart (TCPR-Heart) captures 1,687 heart transplants (1,636 patients) spanning the years 1984-2025. The TCPR-Heart comprises 1,054 transplants with active clinical follow-up: 555 transplants were extracted and abstracted from our modern electronic health record (EHR) in the decade since deployment, providing access to data throughout the patient’s course of heart transplant; 427 were legacy active transplants (transplanted pre-2016 with continued follow-up), and 72 were external transplants (transplanted elsewhere but followed at Methodist).

Additionally, the registry houses a historic cohort of 633 transplants (last follow-up < June 2016) with limited variables. Automated deep phenotyping successfully generated longitudinal data trends across clinical domains, including immunosuppression strategies, rejection, immunologic HLA data, renal function, metabolic profiles, vasculopathy, graft function, hospitalization burden and survival information.

**Conclusion:** This automated framework unifies clinical, administrative, and molecular data streams. By leveraging an automated, regularly updated registry, we established a scalable, high-fidelity data source as a foundation for further innovations and novel applications based on an expertly curated and validated data source.

## INTRODUCTION

The number of heart transplants (HT) performed in the United States has increased significantly in recent years, driven by advances in organ preservation, transportation, and expanded donor availability[1–3]. While short-term outcomes continue to improve, there is a constant need for advancements that can address comorbidities and improve long-term survival after transplantation [4]. Randomized Controlled Trials (RCTs) remain the standard for establishing therapeutic efficacy in medicine yet are often ill-suited for advancement in areas such as heart transplantation, where small sample sizes, high costs and prolonged follow-up requirements limit feasibility. While real-world registries from societies and regulatory bodies have attempted to fill this gap, they are stymied by fragmented data infrastructures and labor-intensive manual abstraction[5], which are susceptible to significant transcription errors [6] and lack granular resolution. Though the United Network for Organ Sharing (UNOS) and the International Society for Heart and Lung Transplantation (ISHLT) registries have made significant contributions, they are not designed to answer hypothesis-driven research questions. [7, 8]

Current availability of clinical data in electronic health record (EHR) creates an opportunity to understand the longitudinal patient journey through deep phenotyping[9] of “real-world” data.[10] The widespread adoption of transplant-specific modules within established EHR interfaces, presents a unique opportunity to modernize this landscape. By leveraging the inherent structure of EHR data, institutions can move towards automated, high-fidelity data management. However, structured data alone is insufficient. A clinically relevant dataset must also ingest unstructured narratives and relevant external data streams and be adapted for specific disease conditions. Finally, while automation offers scale, it has historically been viewed with skepticism regarding data fidelity; thus, any automated framework must include robust validation.

In this study, we describe the methodology used to construct a next-generation, automated, multi-source HT registry at our institution. Housed within a secure, HIPAA-compliant high-performance computing (HPC) environment, this registry integrates automated extraction from our institutional EHR with various external registry data sources. To capture critical clinical nuances trapped in free text, we employed a deterministic, rule-based Natural Language Processing (NLP) engine to convert narrative echocardiography, catheterization, and pathology reports into structured variables, effectively recovering discrete rejection grades and coronary allograft vasculopathy (CAV) classifications otherwise inaccessible to standard queries. We detail the registry’s architecture, data ingestion and validation strategies, and present population-level data, illustrating how automated data engineering enables scalable discovery and quality improvement efforts.

## METHODS

### Study Design and Data Source

This single-center registry was established at the J.C. Walter Jr. Transplant Center at Houston Methodist Hospital (HMH) (Figure 1) and includes all HT patients receiving care between June 2016 and December 2025, identified through our institutional EHR system (Epic Systems, Verona, WI). (IRB Protocol no: Pro00000587).

**Figure 1.**
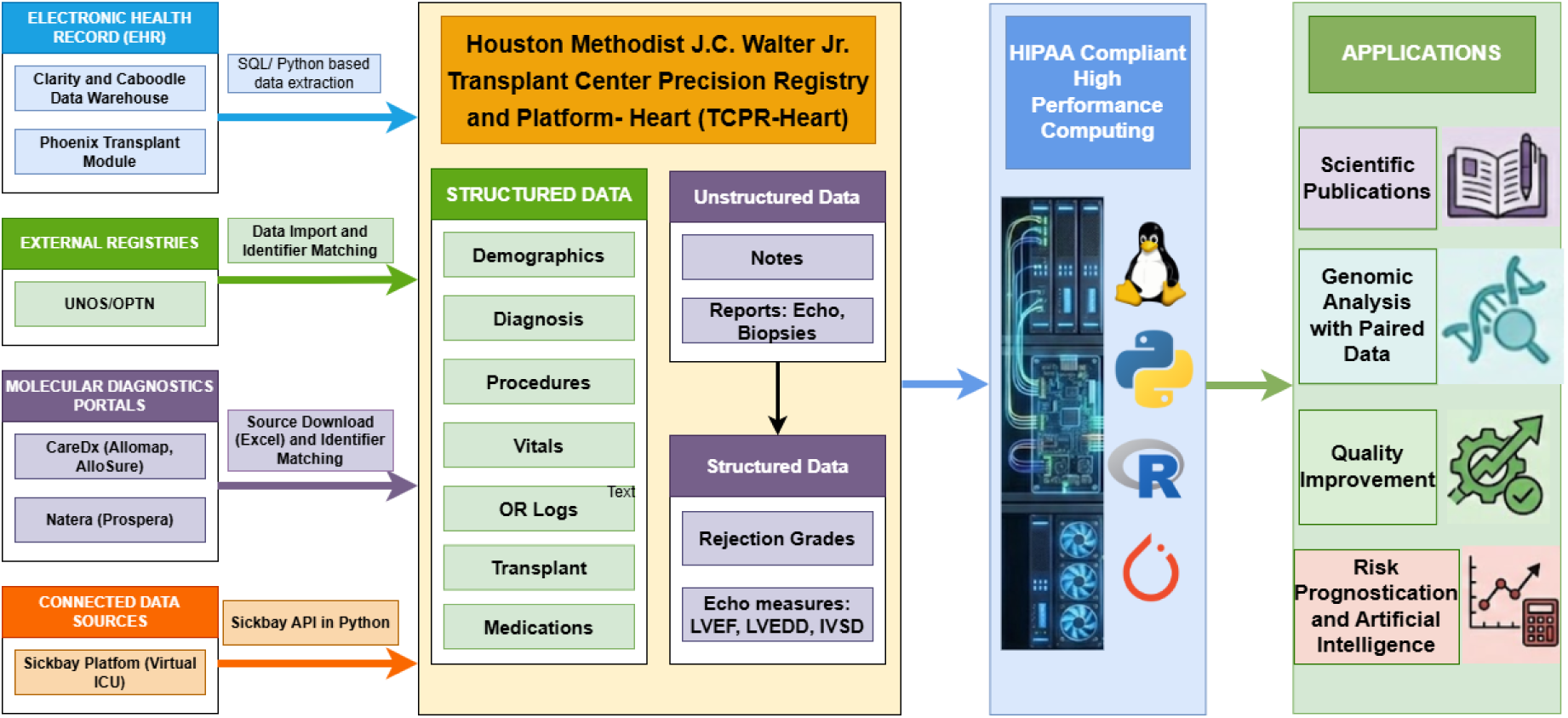
Data Architecture and Workflow of the Automated Multi-Source Heart Transplant Registry. Schematic overview of the data ingestion and processing pipeline used to establish the Houston Methodist J.C. Walter Jr. Transplant Center Precision Registry and Platform-Heart (TCPR-Heart). The infrastructure aggregates data from disparate sources, including the Epic Electronic Health Record (Phoenix Transplant Module and Clarity/Caboodle warehouses), external national registries (UNOS/OPTN) and molecular diagnostic portals (CareDx, Natera). The central repository integrates structured clinical domains with unstructured narrative text (e.g., pathology and echocardiography reports), which undergo deterministic Natural Language Processing (NLP) to recover discrete variables such as rejection grades and hemodynamic measures. Data management and analytics are conducted within a HIPAA-compliant High-Performance Computing (HPC) environment utilizing Python and R, facilitating downstream applications in scientific publication, genomic analysis, quality improvement, and risk prognostication. UNOS: United Network for Organ Sharing; OPTN: Organ Procurement and Transplantation Network; Echo: Echocardiogram HIPAA: Health Insurance Portability and Accountability Act.

### Data Architecture and Infrastructure

Data extraction was anchored on the Epic Phoenix transplant module, which structures the transplant continuum into Referral, Evaluation, Waitlist, and Transplant phases (Supplemental Figure 1). Analysis focused on the ‘Transplant’ phase. Structured data were queried from the EHR data warehouses (Clarity and Caboodle) using structured query language (SQL). Computational tasks were stratified by intensity, with routine processing performed on secure workstations and resource-intensive tasks (e.g., NLP, large-scale merging) executed on a HIPAA-compliant HPC cluster equipped with graphics processing unit (GPUs) and high-memory nodes (Figure 1).

### EHR Data Domains

To ensure comprehensive phenotypic characterization, we extracted a wide array of granular data elements. Extracted domains included medications, laboratory values, vitals, flowsheets, procedural records, demographics, social determinants of health, and unstructured documentation (e.g., clinical notes, imaging, pathology).

### Integration of external molecular diagnostics datasets

Donor-derived cell-free (dd-cf) DNA (AlloSure®, Prospera®) and gene-expression profiling (GEP) (AlloMap®) data were imported from vendor portals as structured files, matched to patient IDs and temporally indexed relative to the transplant date. Risk stratification was applied programmatically using vendor guidelines. For example, AlloSure® levels ≥ 0.20% as “Increased Risk”; AlloMap® scores had time-dependent cutoffs: ≥ 30 for < 6 months post-transplant, and ≥ 34 for > 6 months post-transplant.

### Immunologic dataset

Human Leukocyte Antigen (HLA) laboratory data were curated in a SQL environment from EPIC Clarity, extracting longitudinal Class I and Class II Donor-Specific Antibody (DSA), HLA mismatches and cPRA results. HLA typing was collected for all donor and recipient pairs. High resolution HLA typing was directly obtained from next generation sequencing or was indirectly obtained from typing performed from sequence-specific oligonucleotide probe (SSO) testing. Antibody results from single antigen bead (SAB) testing, antibody levels were collected for all recipients. Donor specific antibodies (DSA) were identified by selecting the individual bead in the SAB array that corresponds to the HLA allele present in the donor genotype. Beads with MFI values over 2000 were considered positive.

### NLP for Unstructured Data

We applied a deterministic, rule-based NLP (using Python-based parsing) pipeline using regular expressions to extract structured variables from echocardiography, coronary angiography, and endomyocardial biopsy reports. Extracted variables included left ventricular dimensions and ejection fraction, rejection grades (0-3R, C4d%), and CAV grades (CAV 0-3). When explicit CAV grades were absent in the report, ISHLT stenosis thresholds were used[11]. Unlike generative large language models, this binary deterministic rule-based NLP either matches a specified pattern or returns null reducing the risk of incorrect values when phrasing changes and instead flags missing values for manual review and code refinement. The custom Regular Expression (Regex) based NLP engine we developed normalizes variable formatting (e.g., handling non-numeric qualifiers), filters physiological outliers, and maps free-text clinical descriptions to standardized phenotypes using extensive lookup dictionaries.

### Outcomes Ascertainment and validation

Mortality was defined as a composite endpoint from EPIC and UNOS/STAR dataset: a patient was classified as deceased if a death date was recorded in either the local Epic EHR or the external UNOS/SRTR dataset. Survival data were obtained from the OPTN Standard Transplant Analysis and Research (STAR) file and merged with local EHR mortality data using deterministic matching based on key identifiers such as Social Security Number (SSN), Donor ID, Recipient First Name, Recipient Last Name, and Recipient Date of Birth (DOB) to create a composite and reliable survival endpoint. To maximize the duration of follow-up, the “last known alive” date was programmatically updated to the latest available timepoint from either source. This was defined as the maximum value between the “last patient encounter” date in Epic and the “last follow-up” date recorded in the UNOS data, thereby minimizing censoring bias. Data integrity was further maintained through automated validation scripts that compared critical fields across sources, such as transplant dates and status between SRTR and Epic. Hospital admissions were identified and categorized using a hierarchical approach. Primary reason for admission was initially derived from billing diagnoses. Script-flagged discrepancies and random subsets of the cohort were periodically reviewed by a transplant data scientist (KP) and a transplant cardiologist (AB) to support high-fidelity ascertainment. NLP-based algorithms were validated by manual review of a sub-cohort of patients for LV dimensions and CAV grading by a transplant cardiologist (AB).

### Data Engineering

Heterogeneity of the source data was managed by engineering a custom, modular software architecture using Python 3.9 and standard Python libraries, including pandas, NumPy, and SQL Alchemy. (Figure 1) Rather than relying on rigid, monolithic queries, we developed a library of domain-specific extraction algorithms with each individual module comprising several hundred lines of specialized code designed to handle the idiosyncrasies of distinct data streams. This programmable infrastructure ensures that the final analytic dataset is not only large in volume but mathematically consistent and reproducible.

### Statistics

Descriptive statistics are presented as medians (IQR) or frequencies (%). Kaplan-Meier methods estimated survival. Longitudinal trends were evaluated at predefined post-transplant intervals. Analyses were performed using R 4.2.1 and Python 3.9 (utilizing the Pandas, SciPy, and Seaborn libraries). Hospitalization burden was calculated as incidence rates per 100 patient-years. A p-value < 0.05 was considered statistically significant.

## RESULTS

### Cohort Description

The registry includes 1,687 HTs (1,636 patients) performed between February 1984 and December 2025. Of these, 1,054 transplants (1,018 patients) had active clinical follow-up extending into the modern EHR era (post June 1, 2016). Table 1 and survival analysis describes the 552 patients (555 transplants) transplanted after full Epic adoption, ensuring comprehensive longitudinal data while the rest of the data description captures the 1054 patients with active follow up into the modern EHR era (Figure 2). An additional 72 patients transplanted at other centers but transferred care to our program were also captured in the registry structure, though they were excluded from the primary outcome analysis to ensure uniform baseline characterization. Median age was 59 years (IQR: 51–65), and 69% were male. The cohort was racially diverse (Table 1). Multi-organ transplants accounted for 35% (Supplemental Figure 2) while 7 (1.3%) were re-transplantations. Dilated Cardiomyopathy was the most common (46%) indication for transplant (Table 1). The dataset also included 14,694 echocardiograms, 1,219 cardiac catheterizations, and 6,027 biopsies, 613 AlloSure® measurements, 550 AlloMap® scores and 1,061 Prospera® assays. Manual resolution was required in <1% of cases.

**Figure 2.**
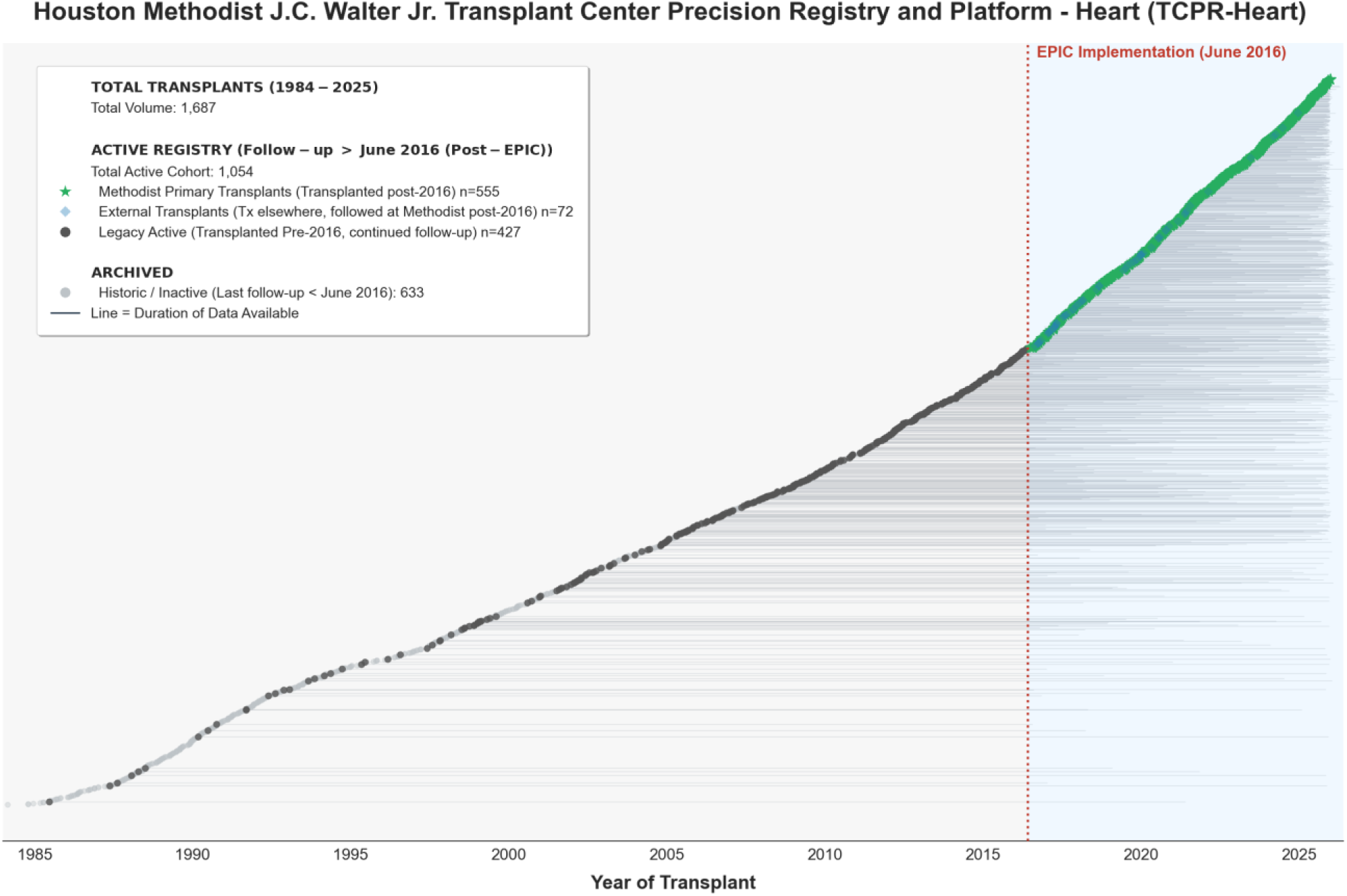
Temporal distribution and cohort structure of the Houston Methodist J.C. Walter Jr. Transplant Center Precision Registry and Platform-Heart (TCPR-Heart) (1984–2025). This longitudinal visualization illustrates the data availability for 1,687 heart transplant records since 1984. The vertical red dotted line denotes the institutional implementation of the Epic Electronic Health Record (EHR) in June 2016, which serves as the inclusion threshold for the deep-phenotyping analysis. Green Stars (n=555) represent the Primary Analytic Cohort reflecting patients transplanted at the center in the post-Epic era, for whom full native granular data is available in our EHR. Blue Diamonds (n=72) represent patients transplanted at other centers but transferred care and managed post-operatively at Houston Methodist in the modern era. Dark Grey Circles (n=427): “Legacy Active” patients transplanted prior to 2016 who continued active clinical follow-up into the modern EHR era. Light Grey Circles (633): Historic cases with no follow-up data in our current EHR extending beyond June 2016. Horizontal lines extending from each marker represent the duration of longitudinal data available within the registry.

**Table 1.** Baseline Demographic, Clinical, and Donor Characteristics of the Primary Analytic Cohort (N = 555 Transplants)

| **Characteristic** | **N = 555<sup>1</sup>** |
| --- | --- |
| **Recipient Age** | 59 (51, 65) |
| **Recipient Sex** |  |
| Female | 172 (31%) |
| Male | 383 (69%) |
| **Recipient Race** |  |
| White, Non-Hispanic | 275 (50%) |
| Black, Non-Hispanic | 171 (31%) |
| Hispanic/Latino | 91 (16%) |
| Asian, Non-Hispanic | 13 (2.3%) |
| Unknown | 3 (0.5%) |
| Amer Ind/Alaska Native, Non-Hispanic | 2 (0.4%) |
| **Primary Diagnosis at Transplant<sup>2</sup>** |  |
| Congenital Heart Disease | 27 (4.9%) |
| Dilated Cardiomyopathy | 253 (46%) |
| Hypertrophic Cardiomyopathy | 10 (1.8%) |
| Ischemic Cardiomyopathy | 168 (30%) |
| Other | 28 (5.0%) |
| Restrictive Cardiomyopathy | 58 (10%) |
| Retransplant/Graft Failure | 7 (1.3%) |
| Valvular Heart Disease | 4 (0.7%) |
| **Recipient Education** |  |
| Associate/Bachelor Degree | 123 (22%) |
| Attended College/Technical School | 151 (27%) |
| Grade School (0-8) | 17 (3.1%) |
| High-School (9-12) or GED | 216 (39%) |
| Post-College Graduate Degree | 44 (7.9%) |
| Unknown | 4 (0.7%) |
| **Recipient Blood Type** |  |
| A | 216 (39%) |
| AB | 23 (4.1%) |
| B | 68 (12%) |
| O | 248 (45%) |
| **Recipient BMI** | 26.2 (23.3, 29.4) |
| **Urgency Status at Listing** |  |
| Adult Status 1 | 19 (3.4%) |
| Adult Status 2 | 324 (58%) |
| Adult Status 3 | 38 (6.8%) |
| Adult Status 4 | 44 (7.9%) |
| Adult Status 5 | 9 (1.6%) |
| Adult Status 6 | 3 (0.5%) |
| Status 1A | 100 (18%) |
| Status 1B | 15 (2.7%) |
| Status 2 | 3 (0.5%) |
| **Waitlist time in days** | 51 (21, 154) |
| **HLA Mismatch Level** |  |
| 1 | 5 (1.0%) |
| 2 | 11 (2.2%) |
| 3 | 55 (11%) |
| 4 | 112 (22%) |
| 5 | 188 (38%) |
| 6 | 127 (26%) |
| **Recipient Diabetes at listing** |  |
| No | 364 (66%) |
| Type I | 4 (0.7%) |
| Type II | 167 (30%) |
| Type Other | 19 (3.4%) |
| Type Unknown | 1 (0.2%) |
| **Recipient Smoking History** | 216 (39%) |
| **Recipient Medical Condition at Transplant** |  |
| Hospitalized Not In Icu | 64 (12%) |
| In Intensive Care Unit | 389 (72%) |
| Not Hospitalized | 84 (16%) |
| **Creatinine at time of Transplant** | 1.30 (1.01, 1.70) |
| **Acute Rejection Episodes Between Transplant and Discharge** |  |
| No | 343 (64%) |
| Yes, at least one episode treated with anti-rejection agent | 142 (26%) |
| Yes, none treated with additional anti-rejection agent | 52 (9.7%) |
| **Length of Stay Post Transplant in Days** | 21 (16, 31) |
| **Previous Solid Organ Transplant** | 17 (3.1%) |
| **Multi-Organ Transplant** | 194 (35%) |
| **Simultaneous Kidney Transplant** |  |
| Left | 34 (6.1%) |
| Right | 83 (14.95%) |
| **Simultaneous Liver Transplant** |  |
| Whole | 46 (8.2%) |
| **Simultaneous Lung Transplant** |  |
| Double | 27 (4.9%) |
| **Heart – Kidney – Liver Transplant** | 4 (0.72%) |
| **Total Ischemic Time** | 3.75 (3.10, 4.20) |
| **Recipient CMV Serostatus** |  |
| Negative | 187 (35%) |
| Positive | 350 (65%) |
| **Recipient EBV Serostatus** |  |
| Negative | 73 (14%) |
| Not Done | 54 (10%) |
| Positive | 406 (76%) |
| Unknown | 4 (0.7%) |
| **Recipient HBV Surface Antibody** |  |
| Negative | 395 (74%) |
| Not Done | 7 (1.3%) |
| Positive | 132 (25%) |
| Unknown | 1 (0.2%) |
| **Recipient HBV Core Antibody** |  |
| Negative | 496 (92%) |
| Not Done | 13 (2.4%) |
| Positive | 28 (5.2%) |
| **Recipient HCV Serostatus** |  |
| Negative | 521 (97%) |
| Not Done | 3 (0.6%) |
| Positive | 13 (2.4%) |
| **Recipient HIV Serostatus** |  |
| Negative | 531 (99%) |
| Not Done | 2 (0.4%) |
| Positive | 4 (0.7%) |
| **Donor Age** | 29 (22, 37) |
| **Donor Sex** |  |
| Female | 162 (29%) |
| Male | 393 (71%) |
| **Donor Race** |  |
| White, Non-Hispanic | 284 (51%) |
| Hispanic/Latino | 152 (27%) |
| Black, Non-Hispanic | 104 (19%) |
| Asian, Non-Hispanic | 6 (1.1%) |
| Amer Ind/Alaska Native, Non-Hispanic | 5 (0.9%) |
| Multiracial, Non-Hispanic | 2 (0.4%) |
| Unknown | 2 (0.4%) |
| **Donor Blood Type** |  |
| A | 97 (17%) |
| A1 | 87 (16%) |
| A2 | 19 (3.4%) |
| A2B | 1 (0.2%) |
| AB | 9 (1.6%) |
| B | 47 (8.5%) |
| O | 295 (53%) |
| **Donor Cause of Death** |  |
| Anoxia | 171 (31%) |
| Cerebrovascular/Stroke | 76 (14%) |
| CNS Tumor | 4 (0.7%) |
| Head Trauma | 292 (53%) |
| Other Specify | 12 (2.2%) |
| **Donor h/o Diabetes** |  |
| No | 523 (94%) |
| Unknown | 7 (1.3%) |
| Yes | 25 (4.5%) |
| **Donor h/o Hypertension** |  |
| No | 471 (85%) |
| Unknown | 9 (1.6%) |
| Yes, >10 Years | 10 (1.8%) |
| Yes, 0-5 Years | 49 (8.8%) |
| Yes, 6-10 Years | 10 (1.8%) |
| Yes, Unknown Duration | 6 (1.1%) |
| **Donor History of Heavy Smoking (>20 Pack Years)** |  |
| No | 544 (98%) |
| Unknown | 6 (1.1%) |
| Yes | 5 (0.9%) |
| **Donor Terminal Total Bilirubin** | 0.80 (0.50, 1.30) |
| **Donor Terminal BUN** | 20 (14, 30) |
| **Donor Terminal Lab Creatinine** | 0.89 (0.69, 1.30) |
| **Donor CMV Serostatus** |  |
| Negative | 154 (28%) |
| Not Done | 6 (1.1%) |
| Positive | 395 (71%) |
| **Donor EBV IgG** |  |
| Indeterminate/E | 5 (0.9%) |
| Negative | 27 (4.9%) |
| Not Done | 11 (2.0%) |
| Positive | 512 (92%) |
| **Donor EBV IgM** |  |
| Negative | 485 (87%) |
| Not Done | 62 (11%) |
| Positive | 8 (1.4%) |
| **Donor HBV Core Antibody** |  |
| Negative | 540 (97%) |
| Positive | 15 (2.7%) |
| **Donor HCV Antibody** |  |
| Negative | 542 (98%) |
| Positive | 13 (2.3%) |
| **Donor HIV Serostatus** |  |
| Negative | 482 (87%) |
| Not Done | 73 (13%) |
| **Donor Toxoplasma IgG** |  |
| Indeterminate/E | 6 (1.2%) |
| Negative | 466 (90%) |
| Not Done | 2 (0.4%) |
| Pending | 1 (0.2%) |
| Positive | 40 (7.8%) |
| **Donor Clinical Infection** |  |
| No | 130 (24%) |
| Unknown | 3 (0.5%) |
| Yes | 414 (76%) |
| **Donor Weight** | 82 (70, 98) |
| **Donor Height** | 174 (168, 180) |
| **Death** | 80 (14%) |

<sup>1</sup>Median (IQR); n (%)
<sup>2</sup>Primary Diagnosis: "Dilated Cardiomyopathy" includes idiopathic, familial, viral, chemotherapy-induced, peripartum, and tachycardia-induced etiologies. "Restrictive Cardiomyopathy" includes amyloidosis, sarcoidosis, and radiation-induced cardiomyopathy. "Other" comprises arrhythmogenic right ventricular dysplasia (ARVD, n=6), pulmonary hypertension (n=4), primary lung pathology (n=3), and COVID-19 ARDS (n=1).

### Immunosuppression

Tacrolimus was used in 100%, 86%, and 84% of patients at Years 1, 3, and 5 post-transplant, respectively. Mycophenolate usage declined from 97% in Year 1 to 66% by Year 5, while Prednisone usage decreased from 97% in Year 1 to 62% by Year 5. mTOR inhibitors utilization was low (Figure 3a). Analysis of a cohort transplanted after 2020 (with data of the first 2 years post-transplant; n=253) highlighted the therapeutic transitions of prednisone wean and early utilization of mTOR inhibitors as impacted by recent clinical protocol changes (Figure 3b).

**Figure 3.**
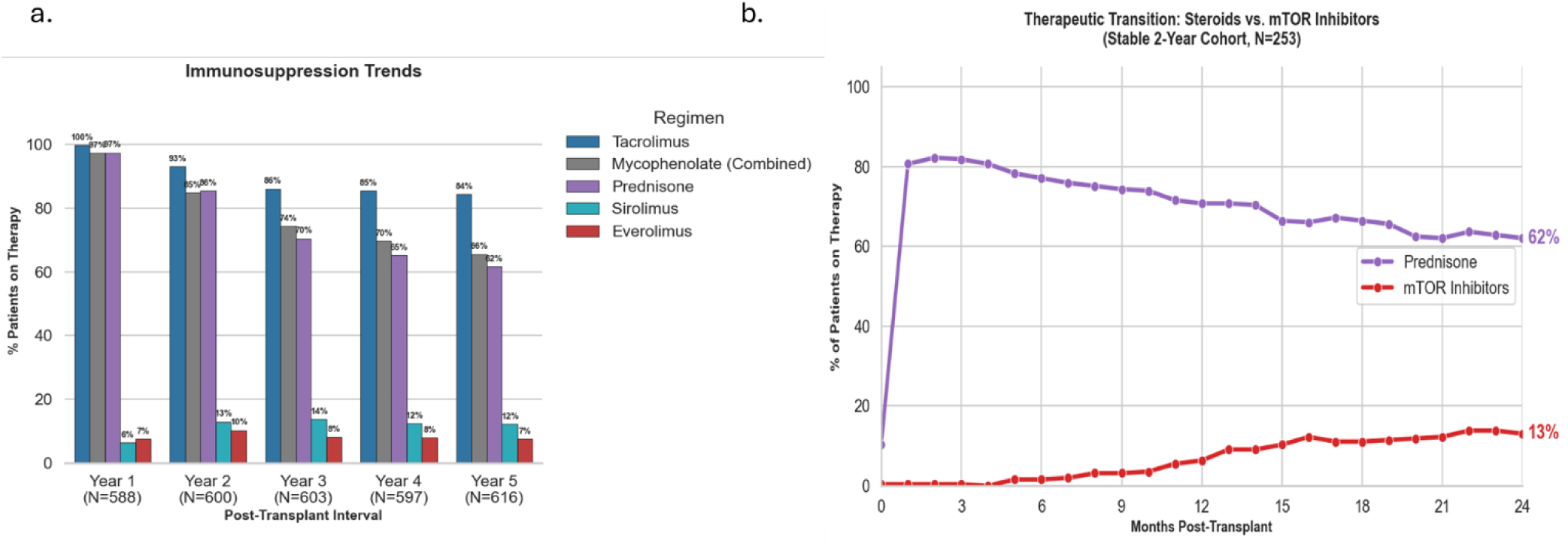
Longitudinal evolution of maintenance immunosuppression regimens. (a) Bar chart illustrating the prevalence of specific immunosuppressive agents (Methodist and External Transplants followed up) at continuous annual intervals from Year 1 through Year 5. Tacrolimus (blue) remains the predominant backbone, utilized in 100% of patients in Year 1 and maintaining 84% prevalence by Year 5. Consistent with weaning protocols, there is a progressive tapering of anti-metabolites (Mycophenolate, grey) from 97% to 66% and corticosteroids (Prednisone, purple) from 97% to 62% over the 5-year period. Data represents period prevalence within each post-transplant year, where Year 1 represents the interval from Day 0 to Day 365. b) Longitudinal line graph of a contemporary sub-cohort (transplanted post-2020, n=253) highlighting the early therapeutic transition over the first 24 months. Visualization captures the increasing utilization of mTOR inhibitors (red line), which rises to 13% by month 24, coinciding with the gradual tapering of Prednisone (purple line) to 62%. This trend reflects the recent implementation of newer institutional immunosuppression protocols for early initiation of mTOR inhibition and weaning of steroids.

### Rejection

NLP analysis of 6,027 endomyocardial biopsy reports, stratified by year identified a 9.1% (547/6,027) overall frequency of Grade 2R rejection. This positivity rate increased from 6.2% (20/320) during 6–12 months post HT to 10.2% (65/637) between years 1–5 and persisting at 7.1% (16/225) in the late post-transplant period (5–10 years) (Supplemental Figure 3a). A C4d immunostaining ≥50% was rare in the early postoperative period (<6 months), occurring in only 1% of biopsies (49/4691). However, the prevalence of high-grade C4d positivity increased over time, rising to 2.2% (7/320) at 6–12 months, 4.2% (27/637) at 1–5 years, and 4.4% (10/225) between 5–10 years (Supplemental Figure 3b). Prospera®, Allomap® and AlloSure® tests showed an increase in the proportion of at-risk patients over time (Figure 4). Of note, we perform surveillance biopsies until month 6. All biopsies performed after that period would be done for either suspicion of rejection or immunosuppression weaning.

**Figure 4:**
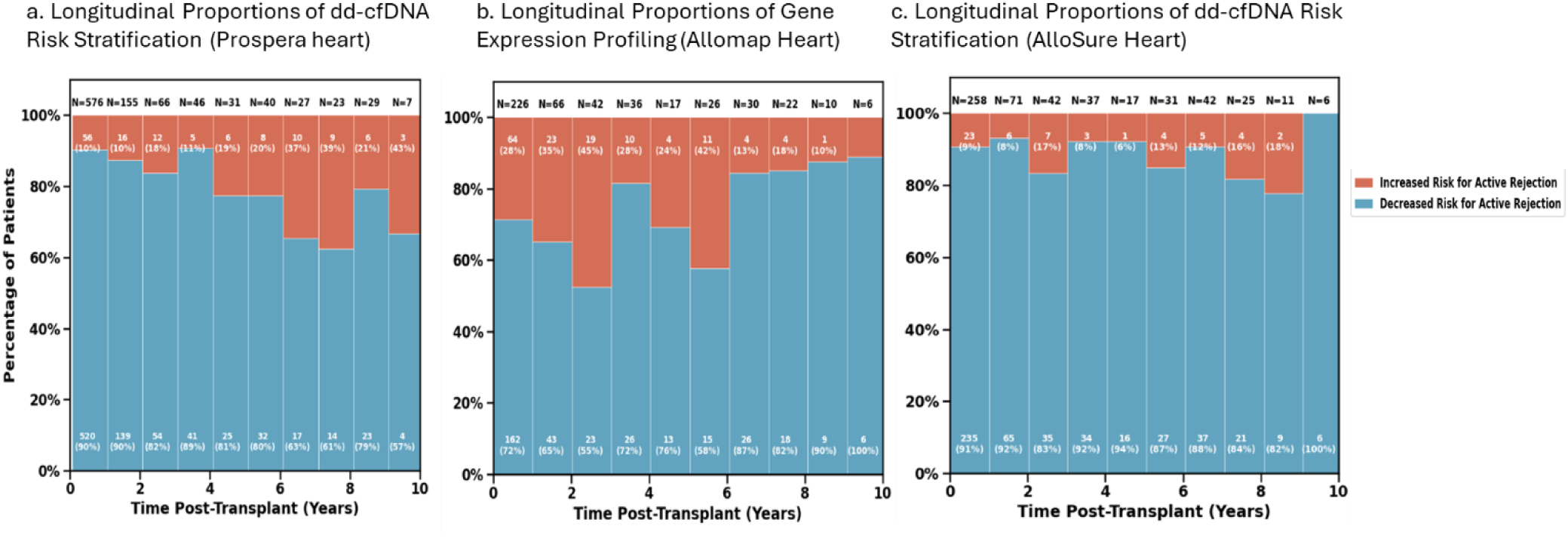
Longitudinal molecular risk stratification using donor-derived cell-free DNA (dd-cfDNA) and gene expression profiling. Comparative stacked bar charts illustrating the annual proportions of “Decreased Risk” (blue) versus “Increased Risk” (red) results over a 10-year post-transplant period for three commercial assays: (a) Prospera Heart (dd-cfDNA), (b) AlloMap Heart (Gene Expression Profiling), and (c) AlloSure Heart (dd-cfDNA). The total number of test results (N) available at each time point is displayed above the bars. Consistent with routine surveillance protocols, the first post-operative year is characterized by a high volume of testing yielding predominantly negative (low risk) results. In subsequent years, a progressive but variable increase in the proportion of “Increased Risk” results is observed across modalities. This data was integrated into the J.C. Walter Jr. Heart Transplant registry from external company data obtained through compliant portals.

### Immunology data

Immunologic characterization was performed via calculated panel reactive antibody (cPRA), number of HLA mismatches, and longitudinal donor specific antibody (DSA) monitoring. The majority had a 0% cPRA (62% for Class I, n=255 and 69% for Class II, n=285; Supplementary Figure 4). HLA mismatches were common with 5-antigen mismatches being the most common (38%, n=188), followed by 6-antigen mismatches (26%, n=127; Table 1). Longitudinal surveillance of DSA burden revealed distinct patterns between Class I and Class II antibodies over the post-transplant course. The test positivity rate for Class I antibodies was 37% (1,356 positive tests) during year 1, remaining relatively stable at 36% (180 positive tests) by year 5, and 37% (511 positive tests) in the long term (≥10y). Conversely, Class II antibody positivity increased progressively, rising from 29% (1,070 positive tests) during year 1 to 44% (198 positive tests) by year 5, and maintaining a high prevalence of 44% (618 positive tests) beyond 10 years (Supplementary Figure 5).

### Renal and Metabolic Outcomes

Median serum creatinine rose from 1.37 mg/dL (IQR: 1.06–1.80) during year 1 (excluding the first 90 days) to 1.82 mg/dL (1.28–3.20) at Year 10. Correspondingly, median estimated Glomerular Filtration Rate (eGFR), calculated using the CKD-EPI 2021 equation[12], declined from 54.9 mL/min/1.73m² during year 1 to 37.4 mL/min/1.73m² by year 10. The burden of renal replacement (defined by dialysis orders, history, or ICD-10 code Z99.2) was assessed as the non-cumulative prevalence within each annual interval. This revealed a U-shaped trajectory: prevalence was 11.3% during year 1 (Day 90–365), dipped to 6.2% at Year 3, rose to 8.4% at Year 5, and reached 11.3% by Year 10. (Supplemental Figure 6)

Pre-transplant diabetes mellitus was present in 34.3% of recipients at the time of listing, with the majority classified as Type II (Table 1). Longitudinal analysis stratified by continuous 1-year intervals (classifying status based on the mean annual value per patient) demonstrated that the proportion of surviving patients with HbA1c levels >6.5% was 18% in the first year (0–1 year interval) to 15% by year 5 (between 4^th^ and 5^th^ year). Abnormal Total Cholesterol levels (>200 mg/dL) were identified in 14%, 10%, and 7% of patients during the 0–1, 2–3, and 4–5-year intervals, respectively. Additionally, the percentage of patients with elevated LDL levels (>100 mg/dL) decreased post-transplant, from 23% during the 0–1 year interval to 15% during the 4–5-year interval. (Figure 5)

**Figure 5:**
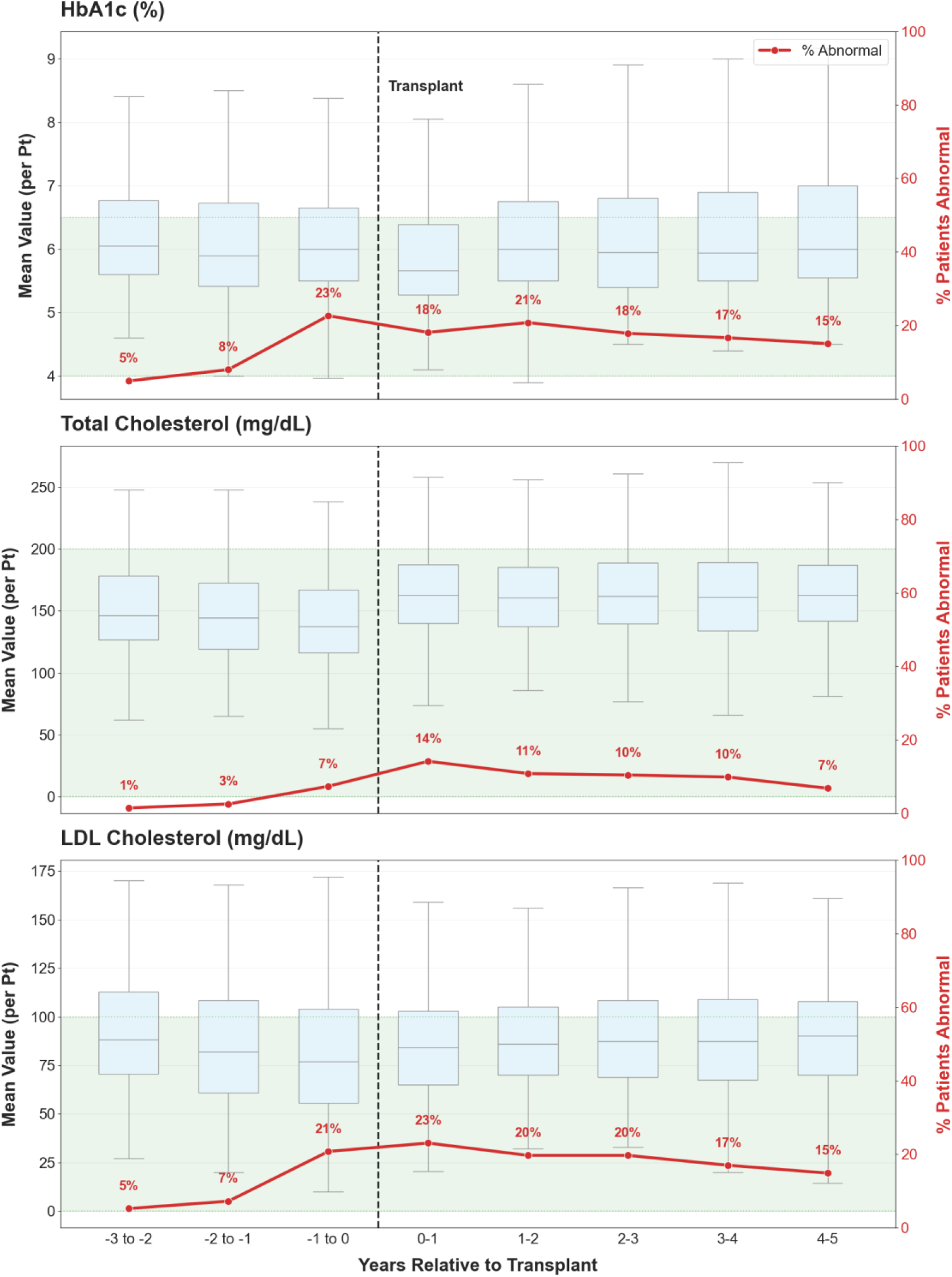
Longitudinal trends in metabolic risk factors pre- and post-transplantation. Composite plot illustrating the trajectory of metabolic control from 3 years prior to 5 years post-transplant. Box-and-whisker plots (left y-axis) display the distribution of values (median, interquartile range) for the cohort at each time interval. The superimposed red line (right y-axis) tracks the percentage of patients exceeding the clinical “Abnormal” threshold. The top panel represents Hemoglobin A1c (HbA1c) (%) as a marker of glycemic control; the proportion of surviving patients with HbA1c >6.5% was 18% in the first year (0–1 year interval) and stabilized at 15% by year 5 (4–5 year interval). The middle panel represents Total Cholesterol (mg/dL), showing a progressive improvement in lipid control, with the prevalence of abnormal levels (>200 mg/dL) declining from 14% in year 1 to 7% by year 5. The bottom panel represents Low Density Lipoprotein (LDL) cholesterol (mg/dL), similarly demonstrating a reduction in the prevalence of elevated LDL (>100 mg/dL) from 23% in the first year to 15% by year 5. The vertical dashed black line denotes the time of transplantation.

### Survival, CAV and Graft function

One, three, and five-year survival rates were 95%, 89% and 84% respectively (Supplemental Figure 7) comparable to national benchmarks. [1] The frequency of Grade 2 CAV was 3.5% at year 5 and increased to 5.7% at 14 years. By 20 years, the frequency of Grade 3 CAV reached 20.0% (Figure 6). Of note, coronary angiograms for CAV surveillance are performed annually starting at year 5 post HT at our center. For validation, grading was compared to manual grading by a transplant cardiologist (AB) in a subset of catheterization reports (n=743, 61% reports) with an agreement rate of 60.1%. The confusion matrix revealed high concordance for Grade 0 (n=287). The distinction between Grade 0 and Grade 1 proved challenging; the algorithm classified 105 Grade 0 cases as Grade 1, and conversely, classified 108 Grade 1 cases as Grade 0. Longitudinal echocardiographic function assessment revealed a ‘waterfall’ distribution of stable functions in the majority of survivors (Figure 7). However, graft dysfunction (defined as LVEF <50% or a drop of >25% of the EF from previous echo) was observed in 159 of 861 patients (18.5%) more than 6 months post-transplant. Among those with dysfunction, 154 met the absolute threshold of LVEF <50%, while 97 experienced a relative decline of >25%. In total, 491 separate dysfunction events were recorded amongst the 159 patients, indicating a burden of persistent or recurrent dysfunction.

**Figure 6:**
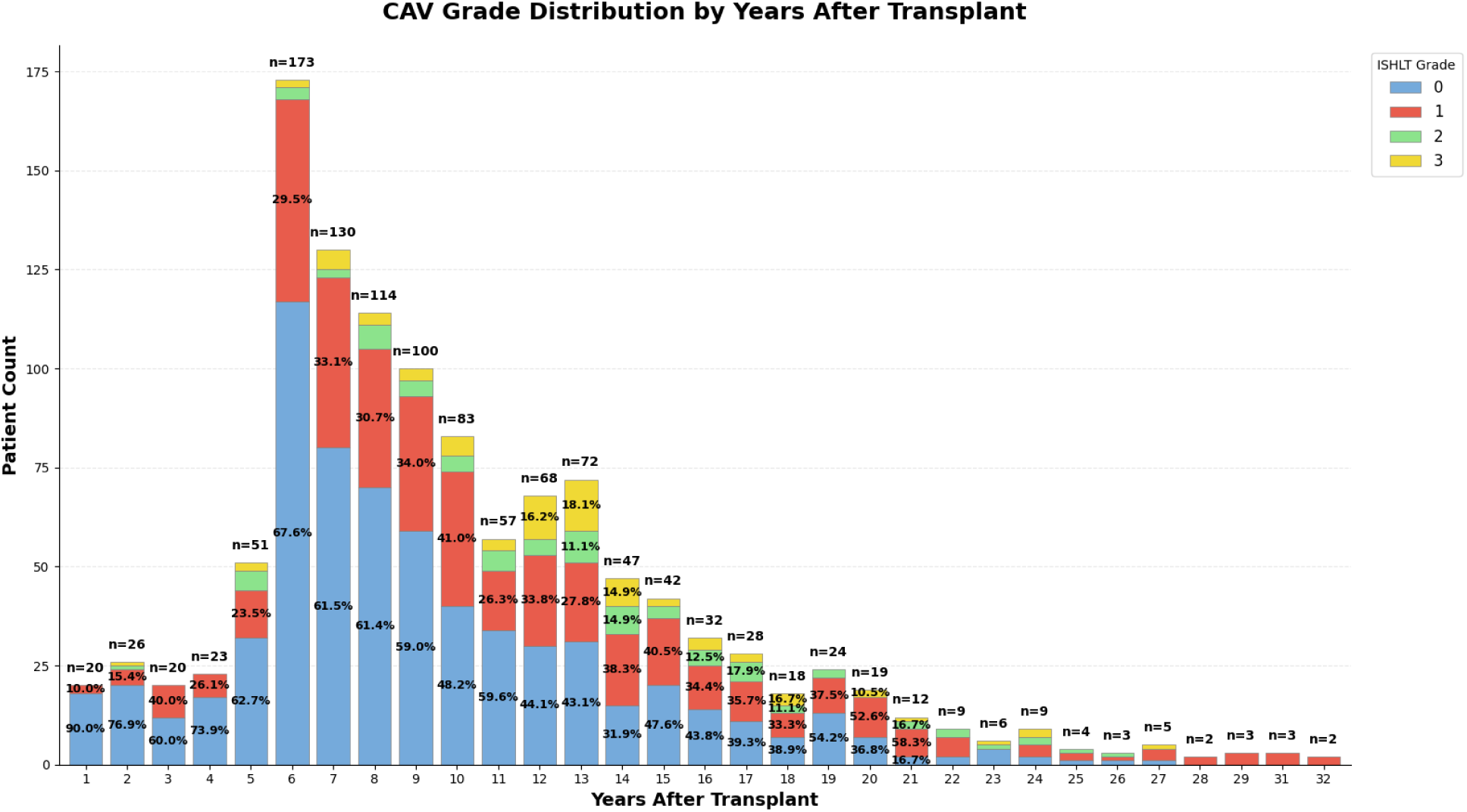
Longitudinal distribution of Cardiac Allograft Vasculopathy (CAV) severity grades. Stacked bar chart illustrates the annual prevalence of CAV according to International Society for Heart and Lung Transplantation (ISHLT) grading. Legend: Grade 0 (No CAV, blue), Grade 1 (Mild, red), Grade 2 (Moderate, green), and Grade 3 (Severe, yellow). Data Volume: The total number of patients with available angiographic grading at each year is indicated above the bars (n=). A distinct increase in data density is observed at Year 6 (5–6 years post-transplant), corresponding with the initiation of routine protocol surveillance angiography. Trend: While Grade 0 (no disease) remains the predominant finding, there is a progressive accumulation of moderate (Grade 2) and severe (Grade 3) vasculopathy in the late post-transplant period (years 10–20).

**Figure 7:**
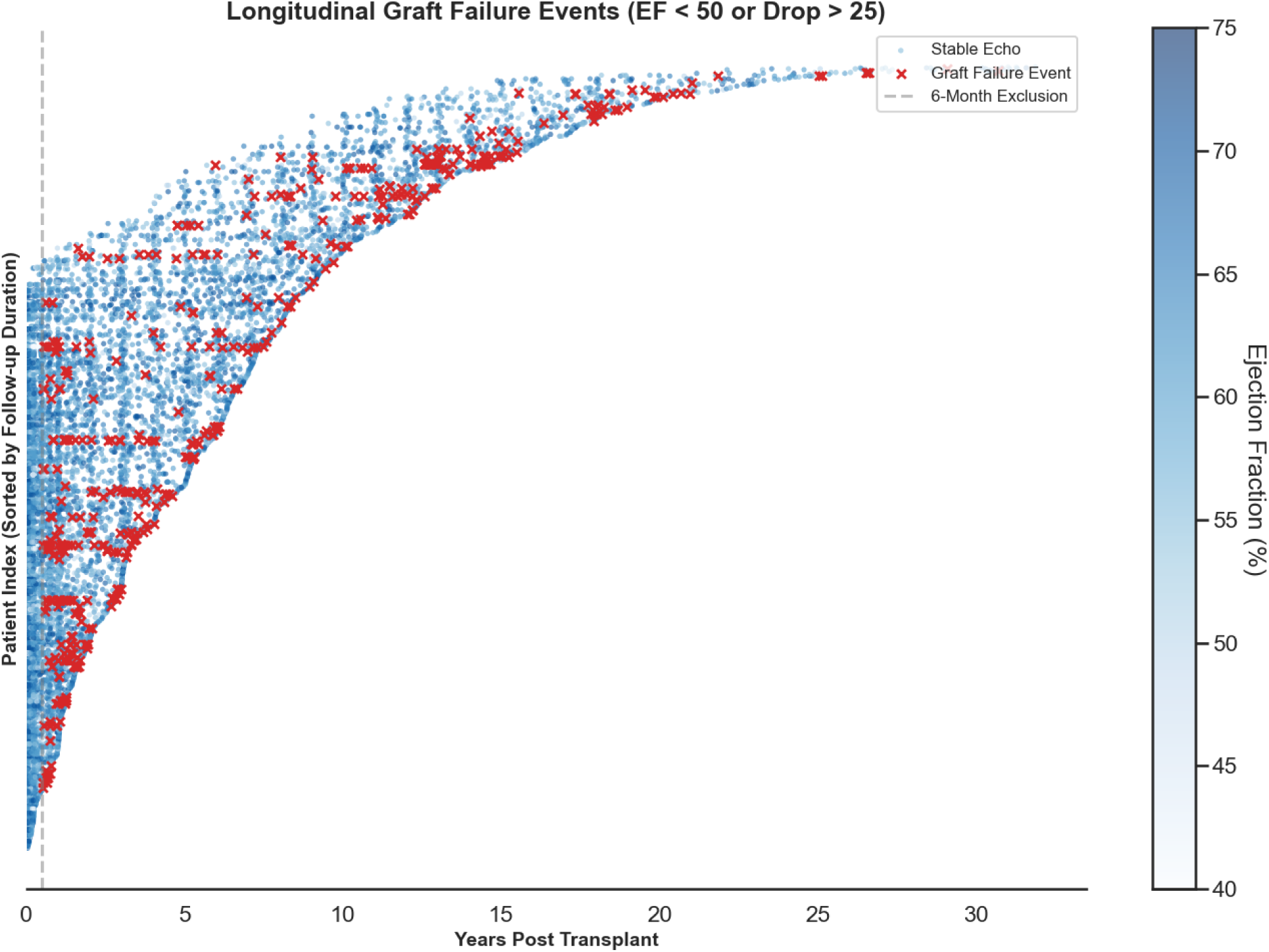
Longitudinal visualization of echocardiographic graft function and dysfunction events. A ‘waterfall’ plot illustrates the longitudinal echocardiographic trajectory of the cohort. Each horizontal line represents a single patient’s clinical course, sorted by total follow-up duration on the y-axis. Blue points represent stable routine surveillance echocardiograms, with color intensity corresponding to Left Ventricular Ejection Fraction (LVEF, %). Red ‘x’ markers denote specific graft dysfunction events. Graft dysfunction was defined as an absolute LVEF <50% or a relative decline of >25% from a previous assessment. The vertical dashed gray line indicates the 6-month post-transplant landmark; events occurring prior to this stabilization period were excluded from the specific graft dysfunction analysis.

### Hospitalizations

In the first year after HT, hospitalization rate was 85.6 admissions per 100 patient-years with a median length of stay (LOS) of 3.7 days (IQR 1.3–7.9). This rate declined to 61.3 in the second year (median LOS 3.0 days; IQR 0.3–6.9) and stabilized at a nadir of ∼ 45.2 admissions per 100 patient-years between years 3 and 5 (median LOS 2.7 days; IQR 0.2–6.4). Notably, the burden of care persisted in the late post-transplant period (years 5–10), with a rate of 53.9 admissions per 100 patient-years (median LOS 3.0 days; IQR 0.3–6.8) (Figure 8).

**Figure 8.**
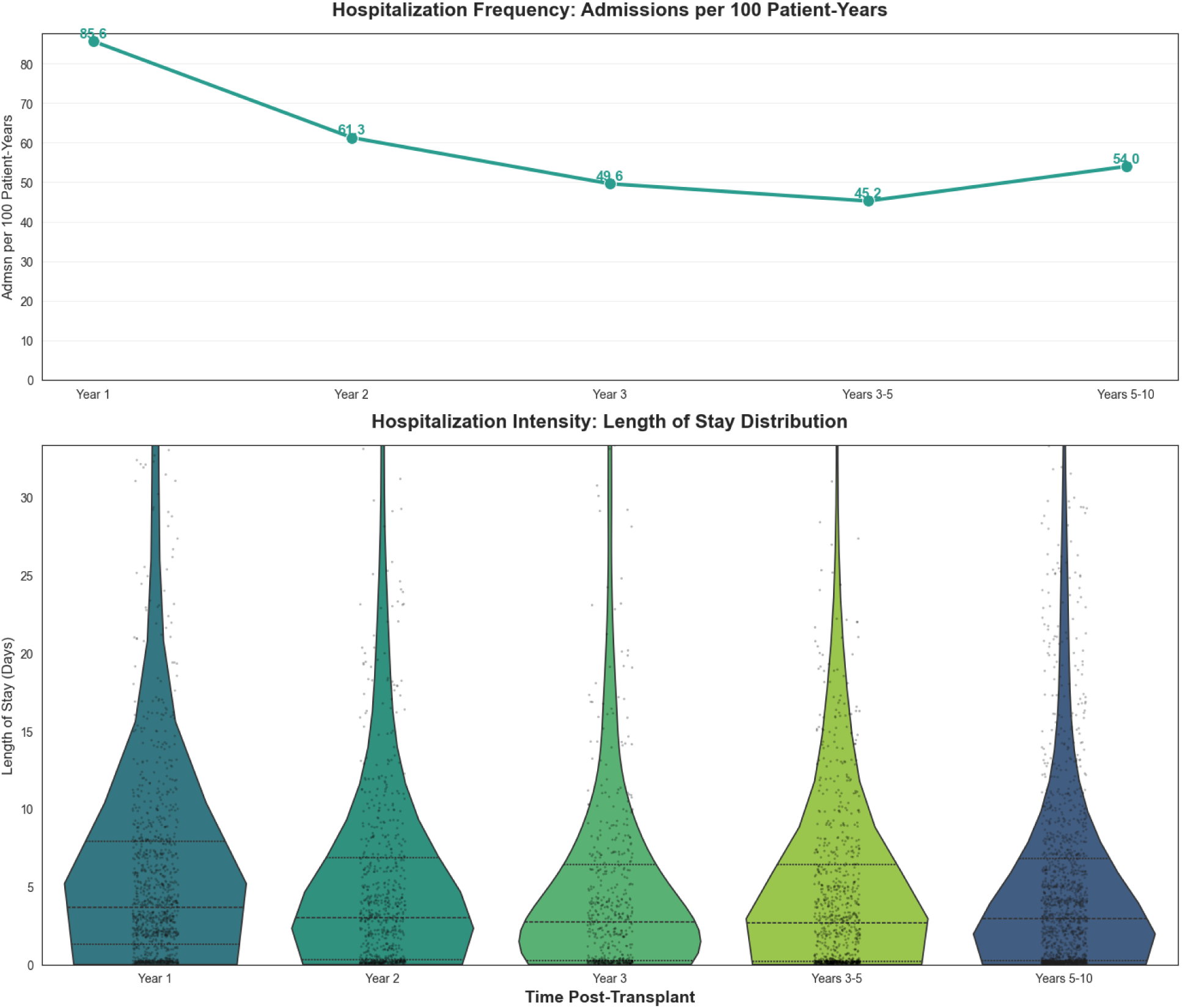
Longitudinal trends in hospital admissions following heart transplantation. Top Panel: Line graph illustrating the incidence density of hospital admissions (events per 100 patient-years). The burden of care is highest in the first post-operative year (85.6 admissions per 100 patient-years), declining to a nadir between years 3–5 (45.2), followed by a persistent burden in the late post-transplant period (54 in years 5–10). Bottom Panel: Violin plots displaying the distribution of Length of Stay (LOS) in days for admissions during each time interval. The width of each plot represents the density of the data, while the internal horizontal dashed lines indicate the median and interquartile ranges. While the frequency of admission declines over time, the median duration of hospitalization remains relatively stable.

## Discussion

Learning from surveillance of the “real-world” of HT patients has historically relied on regulatory and professional society registries. In order to exploit the modernized era of EHR and data science, we present here the creation of the J.C. Walter Jr. Transplant Center Precision Registry and Platform-Heart (TCPR-Heart) data infrastructure created for the heart transplant program and reflective of a high-fidelity medical registry that can overcome many of the limitations of national quality-focused databases.[10]

Leveraging a HPC infrastructure, we can rapidly execute demanding computational tasks (large-scale parsing of unstructured text, sophisticated statistical modeling, machine learning models, and the generation of longitudinal dashboards). By offloading these resource-intensive operations from the real-time SQL environment to the HPC cluster, the registry can continuously consolidate near-real-time data. This data infrastructure currently provides a strong foundation for various research, quality, and operational projects.

Creating an integrated multi-source, automated heart transplant registry represents a transformative approach to addressing data fragmentation in transplant research, overcoming the drudgery of manual data abstraction avoiding errors and conserving human effort. Near real time data access provides enhanced timeliness with frequent updates, and the ability to perform sophisticated longitudinal analyses by consistently capturing data from initial evaluation through long-term follow-up. The reliance on the EHRs SQL-based systems for structured data ensures streamlined, high-quality extraction through SQL Server Management Studio, complemented by deterministic regex-based NLP for precise extraction from narrative clinical documentation.

Our system faces several limitations, including a semi-automated integration that relies on manual data exports from proprietary platforms due to limited API capabilities, potential transcription errors, and variable completeness of historical data before the modern EHR adoption (pre-2016). Although the deterministic NLP approach ensures high accuracy and transparency, it requires regular updates to adapt to evolving documentation practices. Furthermore, the limitations of reliance on subjective clinical reporting are reflected in CAV grading impacted by ‘multiple physician effect’ inherent in historical documentation. This challenge was explicitly demonstrated in our validation phase, where the divergence between NLP-derived grades and a single expert reviewer’s assessment (60.1% agreement) underscores the difficulty in reconciling variable historical grading practices with a standardized, retrospective ‘ground truth’. Imaging based AI applications that can automatically assess coronary lumen patency and left ventricular mass or ejection fraction are being developed and deployed and can be clinically validated utilizing the TCPR-Heart infrastructure. While we were able to successfully import molecular diagnostic tests which are simplistic in reporting, MMDX (molecular microscopic diagnostic system) data architecture needs additional work, which is planned for future iterations. Though, this is a single-center initiative, the custom, modular software architecture was designed to be institution-agnostic providing the technical foundation for multicenter integration within centers using the same EHR. This infrastructure might not be easily replicable in other EHRs.

Our approach uniquely integrates molecular diagnostics and deterministic NLP-driven unstructured data extraction, thereby enhancing the depth and clinical relevance of analyses. Future directions will involve augmenting NLP capabilities with transformer-based machine learning to handle more complex contextual narratives, expanding data modalities (e.g., genomics, advanced imaging), and developing predictive models integrated into clinical decision support systems. This registry also enables expeditious quality improvement by providing near-real time data to assess the impact of protocol changes. Current quality dashboards for most transplant programs focus on publicly reported broad outcomes. Information such as metabolic status, immunosuppression optimization, etc., is not monitored primarily due to limitations of labor-intensive data abstraction. Our data infrastructure enables the creation of such dashboards to provide feedback to programs on intermediate goals that could translate into improved long-term outcomes. For example, as reflected in Figure 3b, the effective implementation of clinical protocols of immunosuppression optimization seems suboptimal compared to our internal goal. Future applications of this data streamlining could be translated into EHR alerts to clinicians. Finally, the ability to conduct robust clinical deep phenotyping creates an infrastructure to build paired genotypic information to advance biological understanding of the complex post-transplant state.

By synergizing high-performance computing with NLP, we have successfully transformed fragmented, unstructured clinical narratives and isolated electronic health records and external data into a unified, high-fidelity research ecosystem. This infrastructure transcends the limitations of traditional, labor-intensive manual registries by enabling the scalable, “deep phenotyping” required to understand the complex longitudinal journey of transplant recipients. The registry established a robust foundation for real-time quality improvement and the generation of “real-world” evidence. Ultimately, this registry serves as a blueprint for modernizing medical and transplant data science by tailoring to a specific population of interest, providing the necessary infrastructure to support predictive analytics, fostering multi-institutional collaboration, and driving the next era of precision medicine in cardiovascular medicine.

In summary, our automated, multi-source heart transplant registry framework addresses key limitations in traditional data collection, providing a robust foundation for longitudinal clinical insights to advance transplant research. Continuous refinements and collaborative expansion efforts with multicenter integration are anticipated to further enhance its utility and generalizability.

### Perspectives

#### Clinical competencies

Clinical care for complex disease states like heart transplantation often is driven by traditional practices stemming from historical institutional protocols. Due to limitations in data, the relatively small number of heart transplants performed across limited number of centers do not provide a high-volume opportunity to learn from real world practices. Further, clinical trials are rarely conducted in such niche populations. Hence the ability to have access to real world data to learn about the impacts of current protocols can provide opportunities to improve patient care and create hypotheses that can be studied further.

#### Translational outlook

By creating a structured, tailored data set that can provide real time data can provide an opportunity for centers to culminate their data and compare impact of the inherent variations of care across many centers in heart transplant care. Furthermore, the ability to generate dashboards for intermediate, process-based outcomes like metabolic control, blood pressure goal, etc provides opportunities for artificial intelligence algorithms to integrate efforts to optimize care through real time alerts in the EHR to physicians and patients.

## Data Availability

Due to HIPAA regulations and patient privacy protections, individual-level data cannot be made publicly available.

## Acknowledgements

None

## Supplementary Figures

**Supplemental Figure 1:**
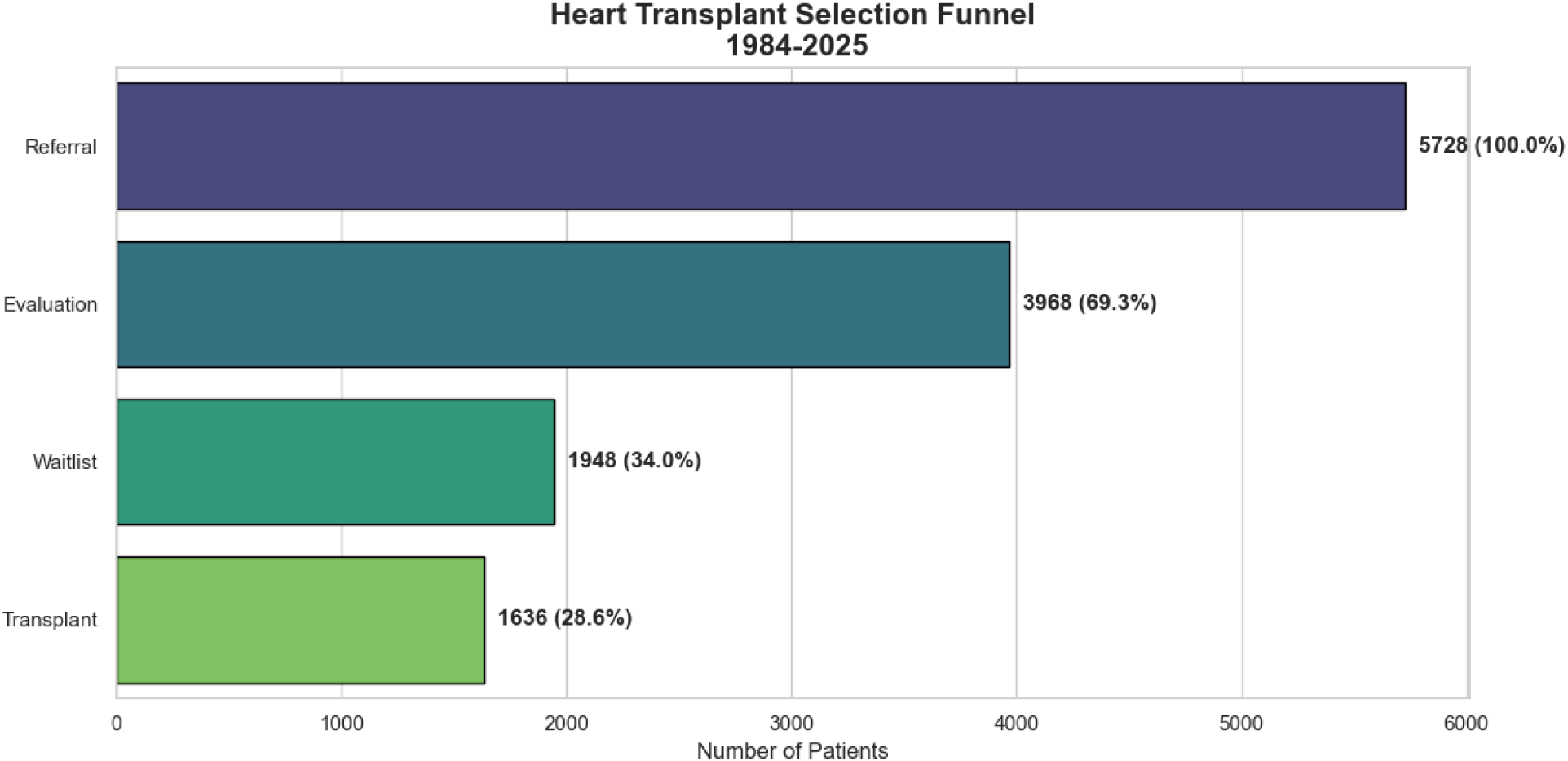
Bar graph representing the episodes of Care segmented in the Epic Phoenix transplant module, showing the number of patients in each phase. These numbers represent years from 1984-2025.

**Supplementary Figure 2.**
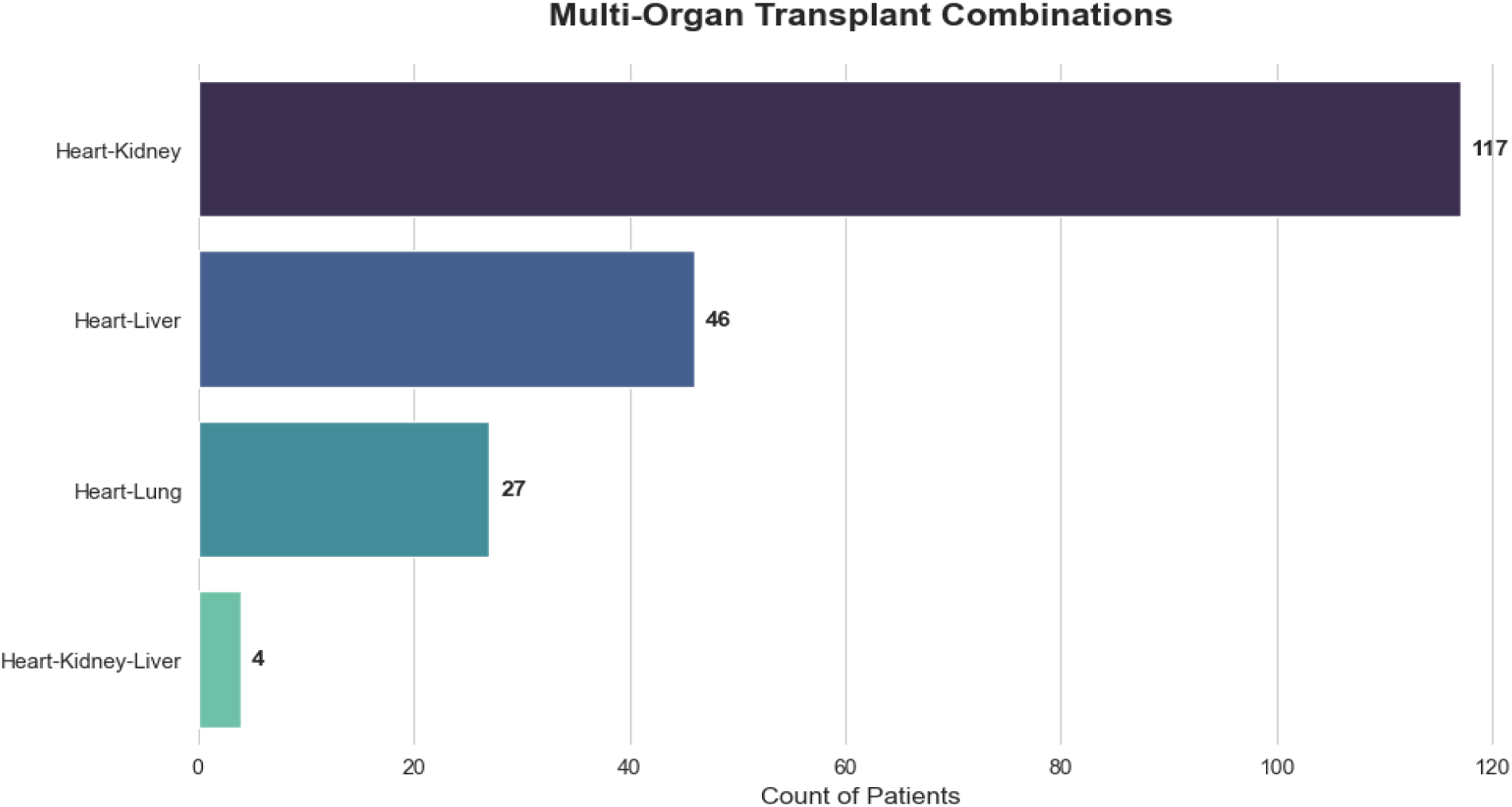
Frequency distribution of multi-organ transplant combinations with heart amongst the modern EHR comprehensive data cohort. Among the 555 heart transplants, 35% (n=194) of recipients underwent multi-organ transplantation. The horizontal bar chart displays the absolute counts for each combination phenotype: Heart-Kidney (n=117) was the most prevalent, followed by Heart-Liver (n=46), Heart-Lung (n=27), and triple-organ Heart-Kidney-Liver (n=4) transplants.

**Supplemental Figure 3:**
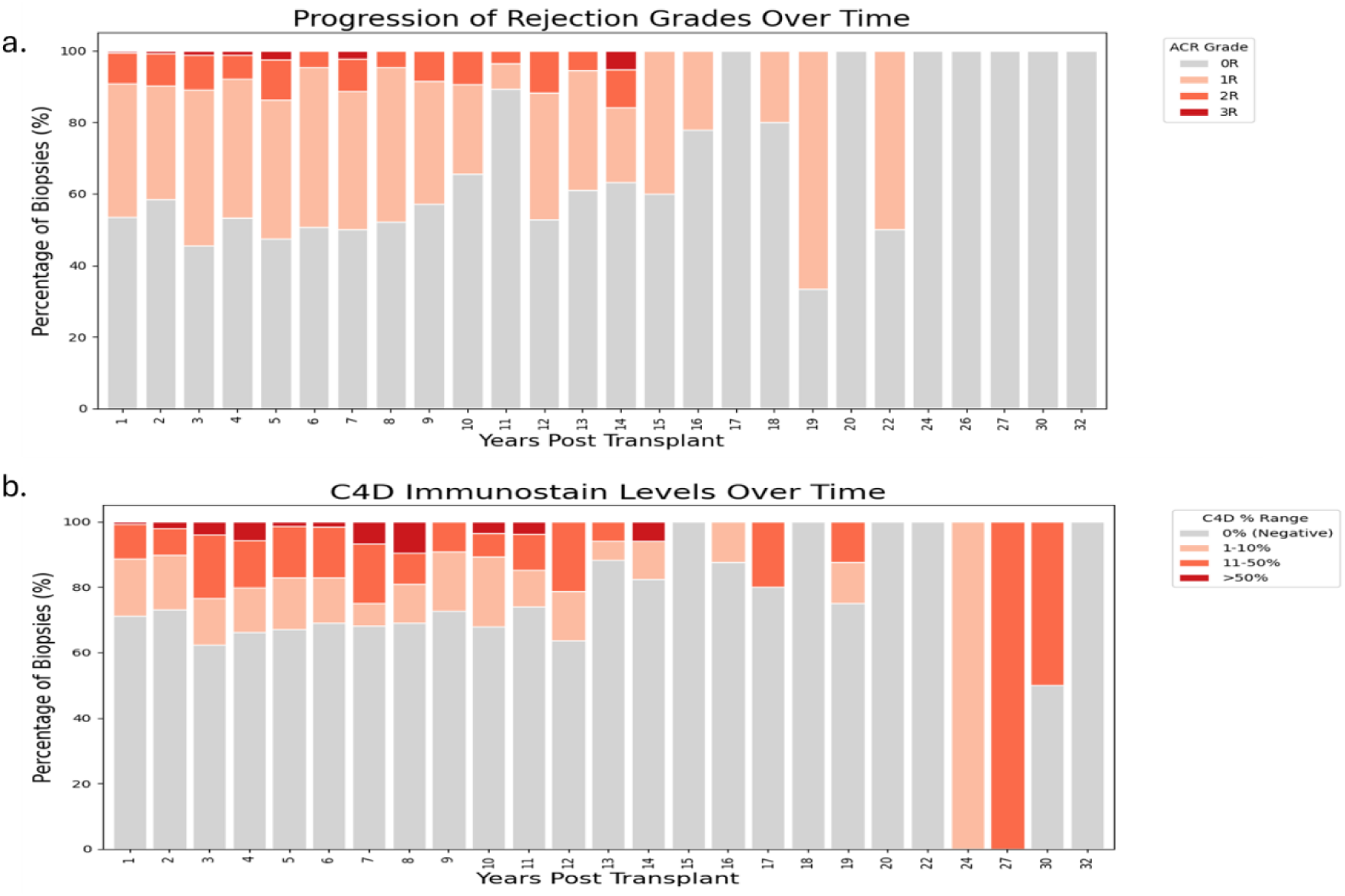
Longitudinal trends in histologic rejection surveillance abstracted using Natural language processing methodology. **(a)** Distribution of Acute Cellular Rejection (ACR) grades over time. Stacked bar chart illustrating the annual proportion of endomyocardial biopsies classified by ISHLT grade. While the majority of biopsies show no rejection (Grade 0R, grey), a persistent burden of moderate (Grade 2R, orange) and severe (Grade 3R, red) rejection events is observed across the post-transplant timeline. **(b)** Antibody-Mediated Rejection (AMR) surveillance reflected by C4d immunostaining. Stacked bars display the percentages of biopsies stratified by C4d staining intensity. High-grade C4d positivity (>50%, dark red) is a pathologic marker for antibody-mediated rejection.

**Supplementary Figure 4.**
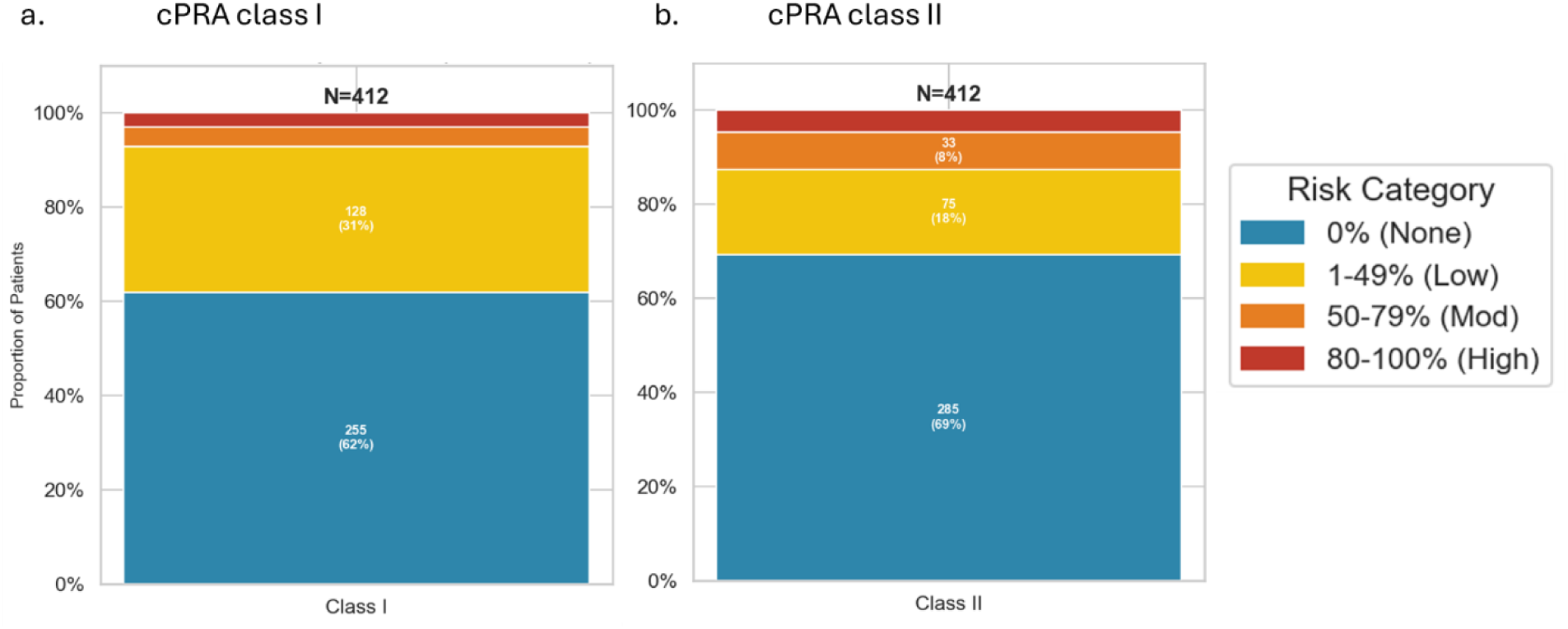
Distribution of baseline sensitization assessed by Calculated Panel Reactive Antibody (cPRA) for **(a)** Class I and **(b)** Class II antigens

**Supplementary Figure 5.**
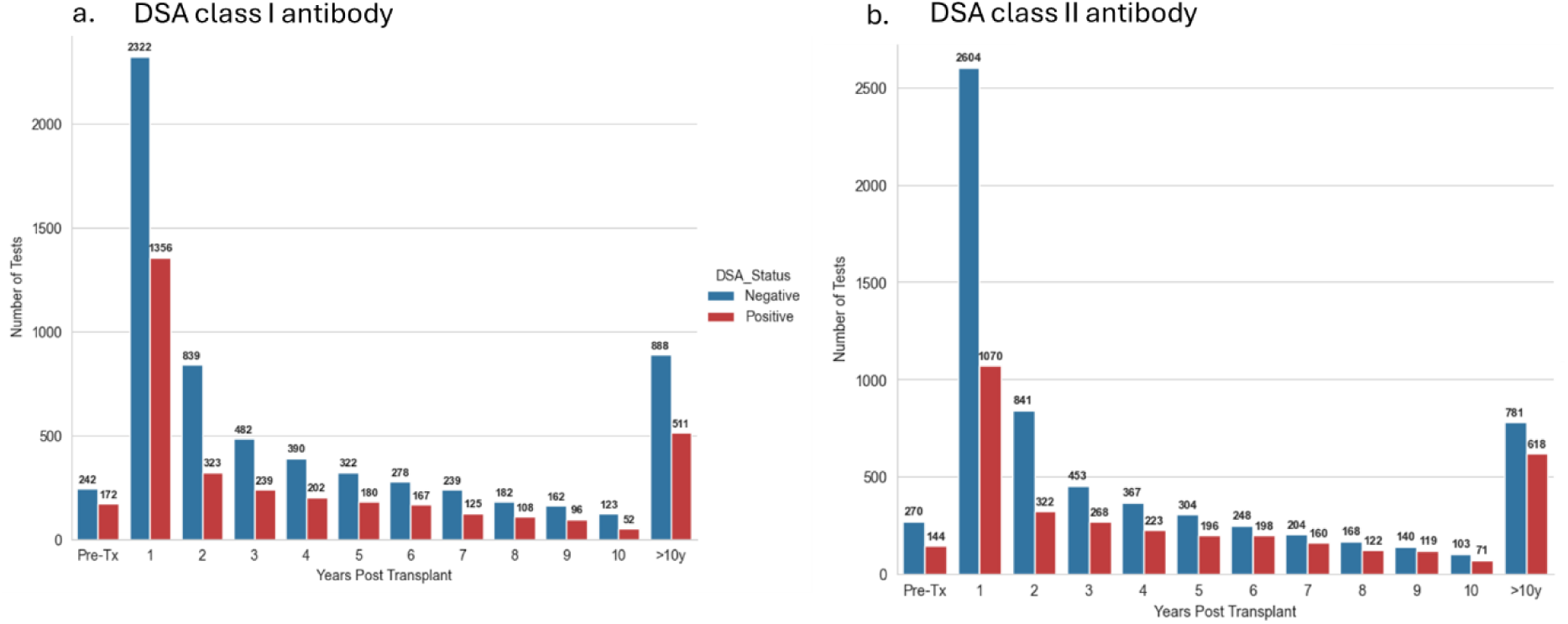
Longitudinal Donor Specific Antibody (DSA) Surveillance. Total burden of immunogenicity assessed by total test volume for **(a)** Class I and **(b)** Class II antibodies at defined post-transplant intervals. Bars represent the absolute number of tests classified as Negative (blue) or Positive (red).

**Supplemental Figure 6:**
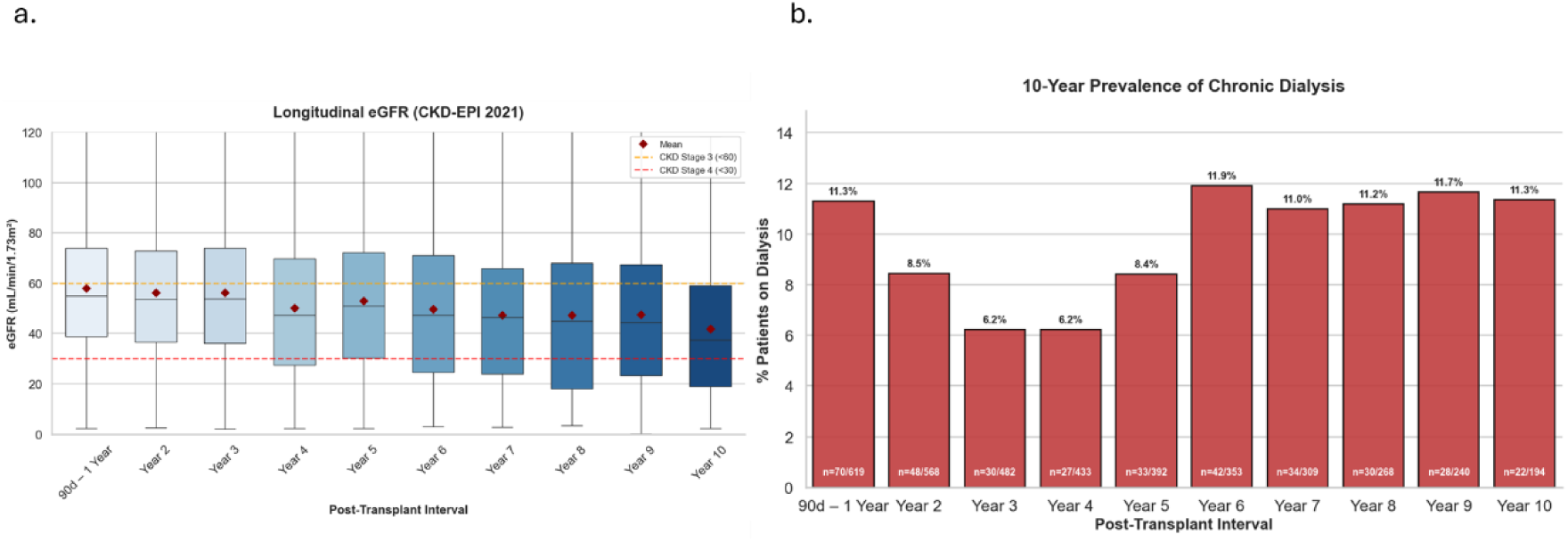
Longitudinal trajectories of renal function and replacement therapy. (a) Distribution of estimated Glomerular Filtration Rate (eGFR). Box-and-whisker plots display eGFR values calculated via the CKD-EPI 2021 equation at continuous annual intervals from Year 1 through Year 10. To exclude acute post-operative fluctuations, Year 1 represents the interval from Day 90 to Day 365. Red diamonds represent the mean eGFR, while colored dashed lines denote Chronic Kidney Disease (CKD) stages: Stage 3 (<60 mL/min/1.73m², orange), Stage 4 (<30 mL/min/1.73m², red), and Stage 5 (<15 mL/min/1.73m², black). The data demonstrates a progressive decline in filtration function, with a median eGFR of 54.9 mL/min/1.73m² at Year 1 decreasing to 37.4 mL/min/1.73m² by Year 10. (b) Prevalence of dialysis dependence. Bar chart illustrating the non-cumulative interval prevalence of surviving patients requiring renal replacement therapy (defined by ICD-10 code Z99.2 or active dialysis orders). The prevalence follows a U-shaped trajectory: an initial rate of 11.3% in the first year (excluding the first 90 days), dipping to a nadir of 6.2% at Year 3, and subsequently rising to 11.3% by Year 10 as chronic allograft nephropathy progresses.

**Supplemental Figure 7:**
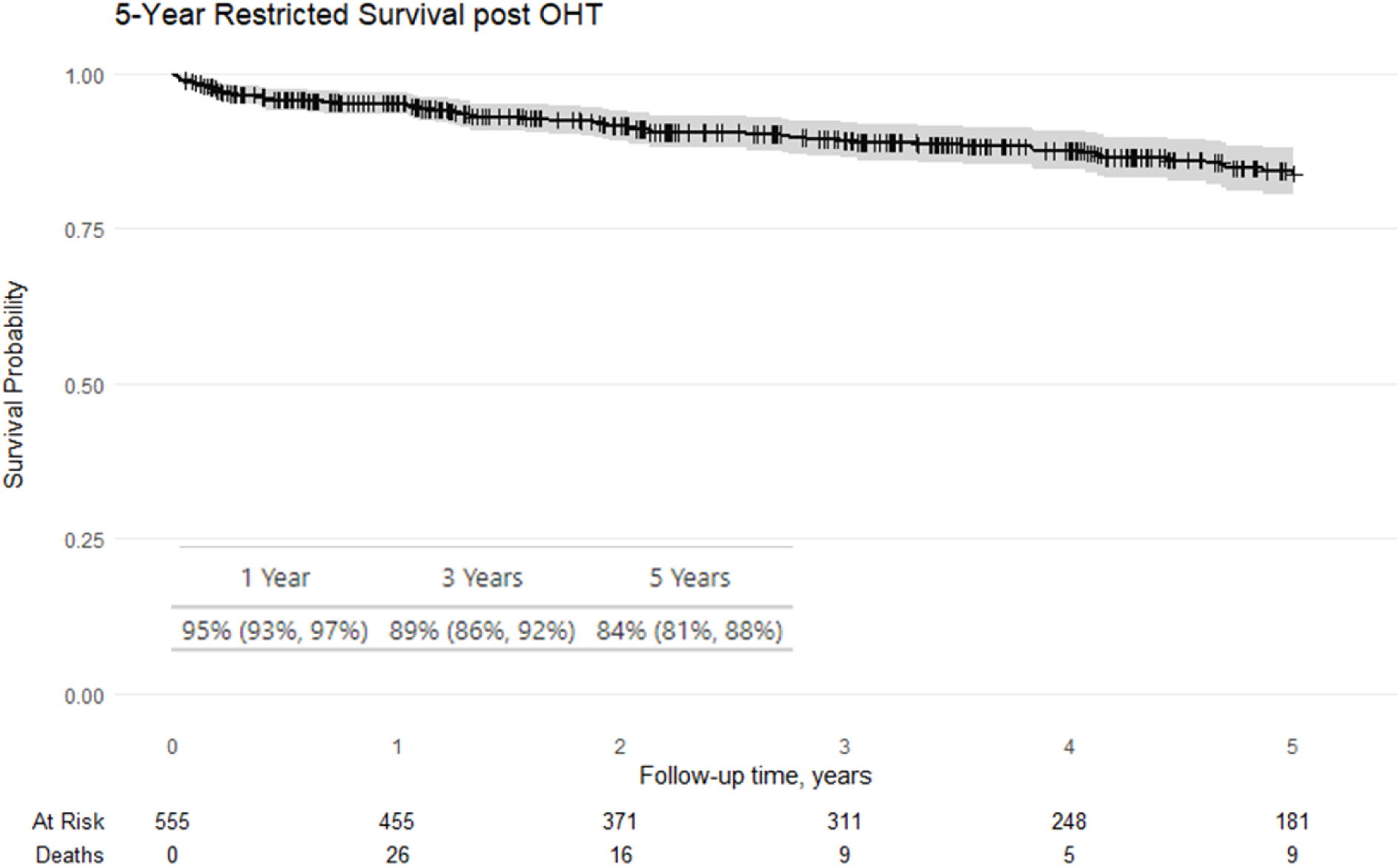
Kaplan-Meier estimates of overall survival in the primary analytic cohort (n=555). Solid black line represents the cumulative survival probability over the first 5 years post-transplantation, with the shaded grey region indicating the 95% confidence interval. Vertical tick marks represent censored individuals. The number of patients at risk and the number of death events at each yearly interval are displayed below the x-axis. Survival rates at 1, 3, and 5 years were 95%, 89%, and 84%, respectively.

